# Endobronchial ultrasound-guided transbronchial mediastinal cryobiopsy: Insights and technical tips from our early experience with 30 patients

**DOI:** 10.1101/2023.08.06.23293719

**Authors:** Chun Ian Soo, Sze Shyang Kho

## Abstract

**Background:** Endobronchial ultrasound-guided transbronchial needle aspiration (EBUS-TBNA) is commonly used to diagnose and stage lung cancer. In real-life practice, limitations are seen with cytology samples from EBUS-TBNA. Obtaining adequate samples can be challenging when faced with necrotic lesions with low cellular yield and the evolving landscape of targeted therapy, necessitating additional samples for comprehensive testing. Hence, transbronchial mediastinal cryobiopsy guided by endobronchial ultrasound (EBUS-TBMC) has emerged as a promising approach for obtaining larger tissue samples. In retrospective review, our aim is to present our early experience regarding the feasibility of performing EBUS-TBMC, employing a similar approach to EBUS-TBNA, followed by the outcomes of our procedures. We include a step-by-step explanation and some recommendations to conduct a successful EBUS-TBMC.

**Method:** Single center retrospective analysis to evaluate the feasibility and utility of EBUS-TBMC cases over six months from July to December 2022.

**Results:** 36 EBUS-TBMC procedures on 30 patients. Moderate sedation was used in 80% of cases. Majority (83.4%) of the patients had biopsy of a single lesion with a median of 3 cryobiopsies (Interquartile range 3-4). The median cryo-activation time was 6 seconds (Interquartile range 6-8). EBUS-TBMC demonstrated a positive yield of 86.1% with an overall diagnostic yield of 83.3%. Mild bleeding occurred in six biopsies (16.7%) which did not required further intervention. No other major complications were observed.

**Conclusion:** EBUS-TBMC is a safe and effective alternative to EBUS-TBNA. Histology samples obtained through EBUS-TBMC have the potential to increase confidence in diagnosing and staging lung cancer, thereby alleviating concerns about tissue inadequacy.

## 1. Introduction

Endobronchial ultrasound-guided transbronchial needle aspiration (EBUS-TBNA) is a minimally invasive technique commonly employed for diagnosing and staging lung cancer [1-2]. However, diagnostic challenges of EBUS-TBNA are seen when dealing with low-cellularity lesions, particularly those involving necrosis or non-malignant conditions [3]. With the emergence of targeted therapy in non-small cell lung carcinoma (NSCLC), additional samples are necessary for immunological profiling and molecular testing. Although transbronchial forceps biopsy and larger cytology needles have been attempted in such cases, their efficacy remains uncertain [3].

A newer technique, transbronchial mediastinal cryobiopsy guided by endobronchial ultrasound (EBUS-TBMC), has recently gained popularity for obtaining larger tissue samples. Despite its growing use, the feasibility, optimal sampling strategy, diagnostic accuracy, and safety profile of EBUS-TBMC are subjects of ongoing debate. As regular EBUS-endoscopists, we have also explored EBUS-TBMC. In this short commentary, we share our early experience with the feasibility and utility of EBUS-TBMC. In addition, we offer a detailed exposition of the employed technique, with the intent of enriching readers’ comprehension and offering invaluable insights into the procedure and its clinical applications.

## 2. Methods

We conducted a retrospective analysis of EBUS-TBMC cases performed at a single tertiary center between July and December 2022. The clinical data of the patients, including radiological images and EBUS-TBMC findings were traced using the hospital’s electronic medical records. The authors had access to participants information during data collection.

An experienced operator carried out the EBUS-TBMC procedure using a 1.1-mm flexible cryoprobe (Erbecryo 20402-401, Tubingen, Germany). The procedure was conducted under either moderate sedation or general anesthesia, following established guidelines [4]. The entry point and biopsy track (BT) for the cryoprobe were created using a needle (19, 21, or 22-gauge), followed by an unspecified number of cryo-activations per biopsy. The choice of needle size was based on the operator’s assessment of the type and location of the lesion, while the number of cryo-activations per biopsy was determined at the operator’s discretion. The frozen biopsy tissue was retracted en-bloc with the scope and probe, and the thawed samples were fixed in formalin. Rapid onsite cytology examination was not available during the procedures. For our analysis, we defined a positive yield as a definitive histology diagnosis from each biopsy, while the diagnostic yield was based on diagnoses from available histology samples. The study protocol received approval from the Medical Research and Ethics Committee, Ministry of Health Malaysia (NMRR ID-23-00577-6XP), and informed consent was waived for this retrospective study.

## 3. Results

A total of 36 EBUS-TBMC procedures were performed on 30 patients. Moderate sedation was utilized in 80% of cases. On average, 3 EBUS-TBMCs were conducted per patient (interquartile range [IQR]: 3-4), with an activation time of 6 seconds (IQR: 6-8). In the majority of cases, EBUS-TBMC was performed on a single lesion, accounting for 83.4% (25 patients) of the total. Among the biopsies conducted, 83.3% (30 biopsies) targeted lymph nodes, while the remaining 16.7% (6 biopsies) were performed on masses. Only two instances of failure in cryoprobe insertion were observed.

The median cumulative tissue size retrieved during EBUS-TBMC procedures was 6 mm (IQR: 5-8). Notably, the positive yield of EBUS-TBMC was 86.1% (31 lesions), contributing to an overall diagnostic yield of 83.3% (25 cases). Table 1 presents the baseline characteristics and diagnostic yield. Mild bleeding occurred in six cases (16.7%) but did not require any intervention. Fortunately, no pneumothoraces, pneumomediastinum, mediastinitis, or other complications were observed.

**Table 1.**
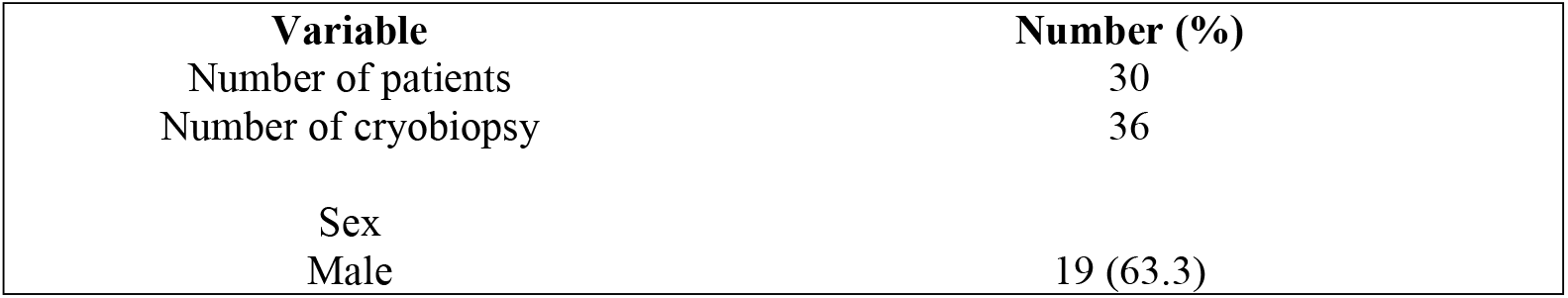
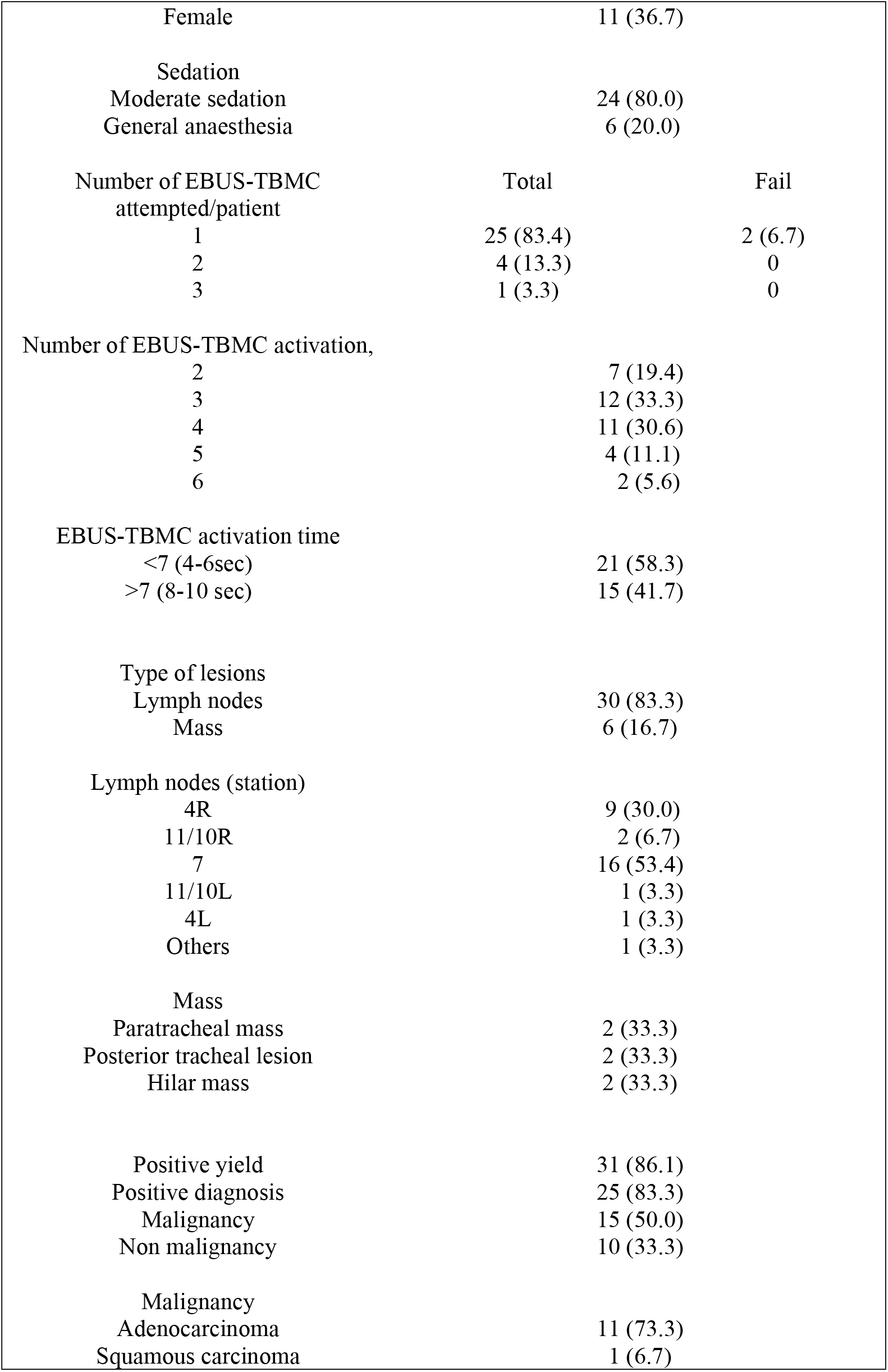
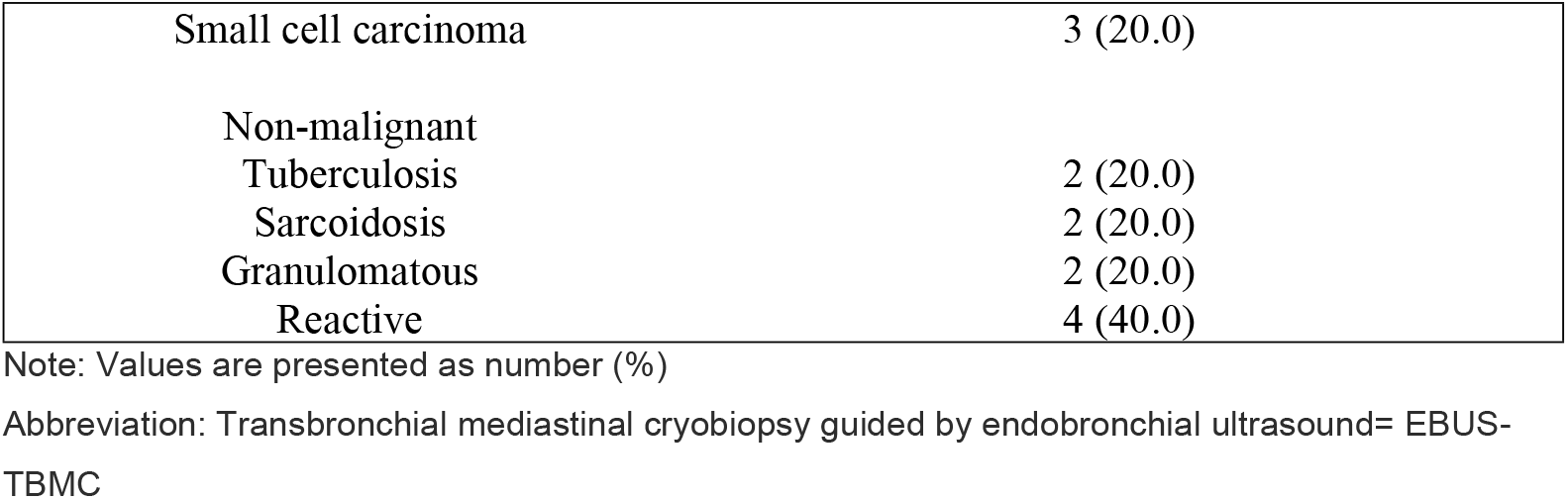
Baseline characteristics and diagnostic yield of EBUS-TBMC.

| Variable | Number (%) |
| --- | --- |
| Number of patients | 30 |
| Number of cryobiopsy | 36 |
| Sex |  |
| Male | 19 (63.3) |

|  |  |  |
| --- | --- | --- |
| Female | 11 (36.7) |  |
| Sedation |  |  |
| Moderate sedation | 24 (80.0) |  |
| General anaesthesia | 6 (20.0) |  |
| Number of EBUS-TBMC attempted/patient | Total | Fail |
| 1 | 25 (83.4) | 2 (6.7) |
| 2 | 4 (13.3) | 0 |
| 3 | 1 (3.3) | 0 |
| Number of EBUS-TBMC activation, |  |  |
| 2 | 7 (19.4) |  |
| 3 | 12 (33.3) |  |
| 4 | 11 (30.6) |  |
| 5 | 4 (11.1) |  |
| 6 | 2 (5.6) |  |
| EBUS-TBMC activation time |  |  |
| <7 (4-6sec) | 21 (58.3) |  |
| >7 (8-10 sec) | 15 (41.7) |  |
| Type of lesions |  |  |
| Lymph nodes | 30 (83.3) |  |
| Mass | 6 (16.7) |  |
| Lymph nodes (station) |  |  |
| 4R | 9 (30.0) |  |
| 11/10R | 2 (6.7) |  |
| 7 | 16 (53.4) |  |
| 11/10L | 1 (3.3) |  |
| 4L | 1 (3.3) |  |
| Others | 1 (3.3) |  |
| Mass |  |  |
| Paratracheal mass | 2 (33.3) |  |
| Posterior tracheal lesion | 2 (33.3) |  |
| Hilar mass | 2 (33.3) |  |
| Positive yield | 31 (86.1) |  |
| Positive diagnosis | 25 (83.3) |  |
| Malignancy | 15 (50.0) |  |
| Non malignancy | 10 (33.3) |  |
| Malignancy |  |  |
| Adenocarcinoma | 11 (73.3) |  |
| Squamous carcinoma | 1 (6.7) |  |
| Small cell carcinoma | 3 (20.0) |
| Non-malignant |  |
| Tuberculosis | 2 (20.0) |
| Sarcoidosis | 2 (20.0) |
| Granulomatous | 2 (20.0) |
| Reactive | 4 (40.0) |

Note: Values are presented as number (%)
Abbreviation: Transbronchial mediastinal cryobiopsy guided by endobronchial ultrasound= EBUS-TBMC

## 4. Discussion

### 4.1 Technical aspect

EBUS-TBMC, akin to EBUS-TBNA, can be carried out with moderate sedation [5-6]. For enhanced patient tolerance or when dealing with small or multiple lesions, the procedure is preferably performed under deep sedation or general anaesthesia [7]. In our study, we primarily conducted EBUS-TBMC under moderate sedation, as regular anaesthesia support was unavailable.

When EBUS-TBMC is employed as an ancillary test for EBUS-TBNA, it is advisable to conduct EBUS-TBMC as the initial procedure. This approach allows for the precise identification of a proper and consistent entry point and BT created by repeated punctures of the TBNA needle. In our procedures, we did not use high-frequency needle knives, in line with a previous studies [6]. However, using a needle knife may potentially reduce the overall procedural duration [5-6].Using a 21-gauge needle, around 5-6 agitations over the capsule than the cortex, following a similar trajectory visible on ultrasound and bronchoscopic vision, are required to create a BT after puncturing the membranous part of the tracheobronchial wall. The number of agitations may need to be adjusted depending on whether a larger or smaller needle is used. The cryoprobe is then inserted precisely perpendicular to the BT into the lesion.

EBUS-TBMC can be conveniently performed at lymph node stations commonly sampled via EBUS-TBNA. However, our experience reveals that EBUS-TBMC presents challenges for posterior tracheal lesions due to the difficulty of rotating the scope 180 degrees. Nevertheless, we believe that mastering EBUS-TBMC requires a steep learning curve. Fig 1 provides a pictorial narrative of the procedure, and Fig 2 depicts the landmarks corresponding to the complexity level of performing EBUS-TBMC.

**Fig 1:**
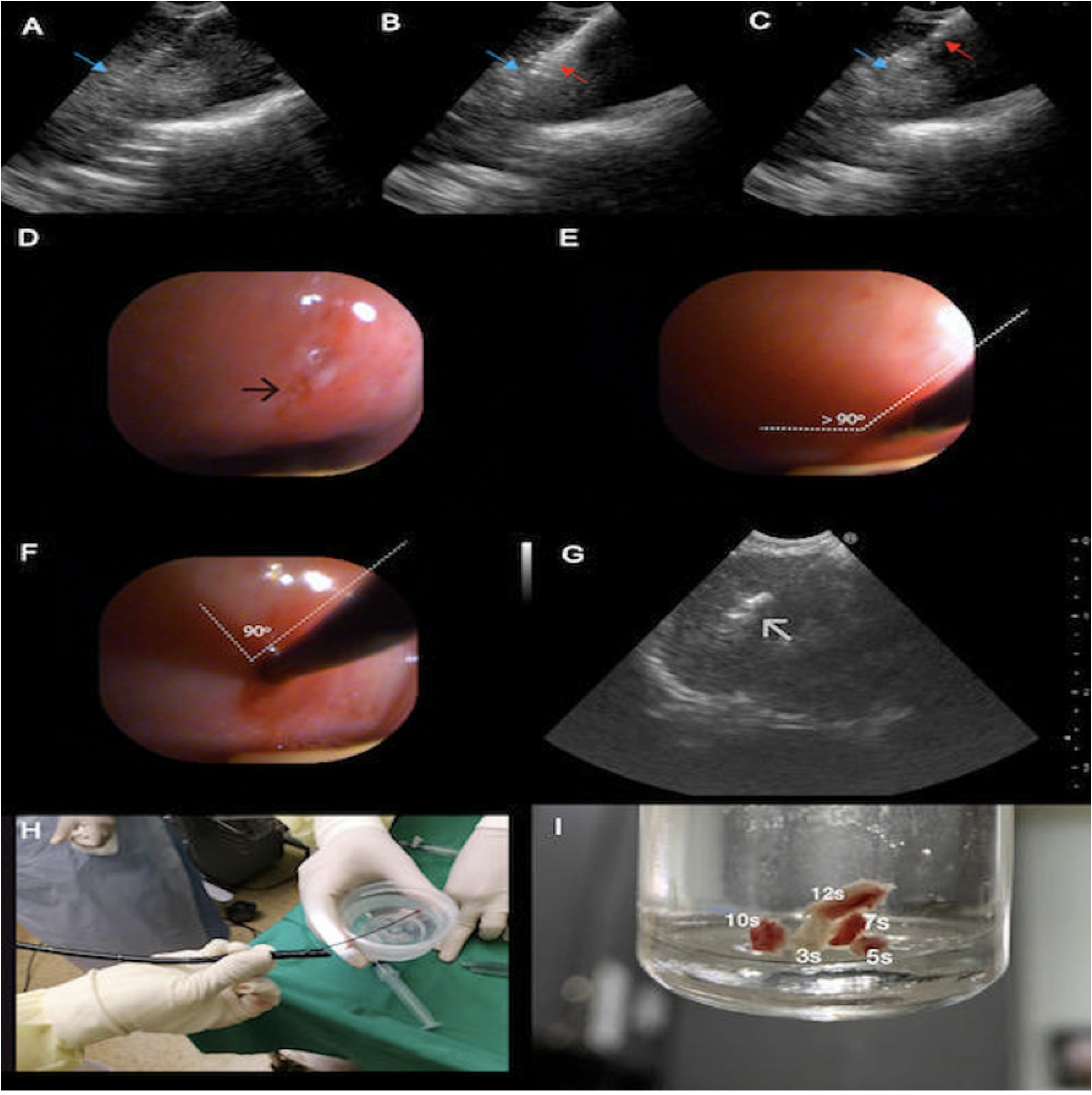
Pictorial narrative of the EBUS-TBMC procedure (In alphabetical sequence) (A) Biopsy track created by TBNA needle seen on ultrasound (blue arrow), (B and C) Biopsy track (blue arrow) and TBNA needle (red arrow) seen at the same trajectory and different depth, (D) Entry point seen after a 22 gauge TBNA needle puncture (black arrow), (E) Avoiding entry of cryoprobe at obtuse angle, (F) Maintain perpendicular entry of cryoprobe, (G) Ensure cryoprobe is clearly visible in lesion, (H) Samples thawed in saline and fixed in formalin, (I) Sample size obtained at different cryo-activation time.

**Fig 2:**
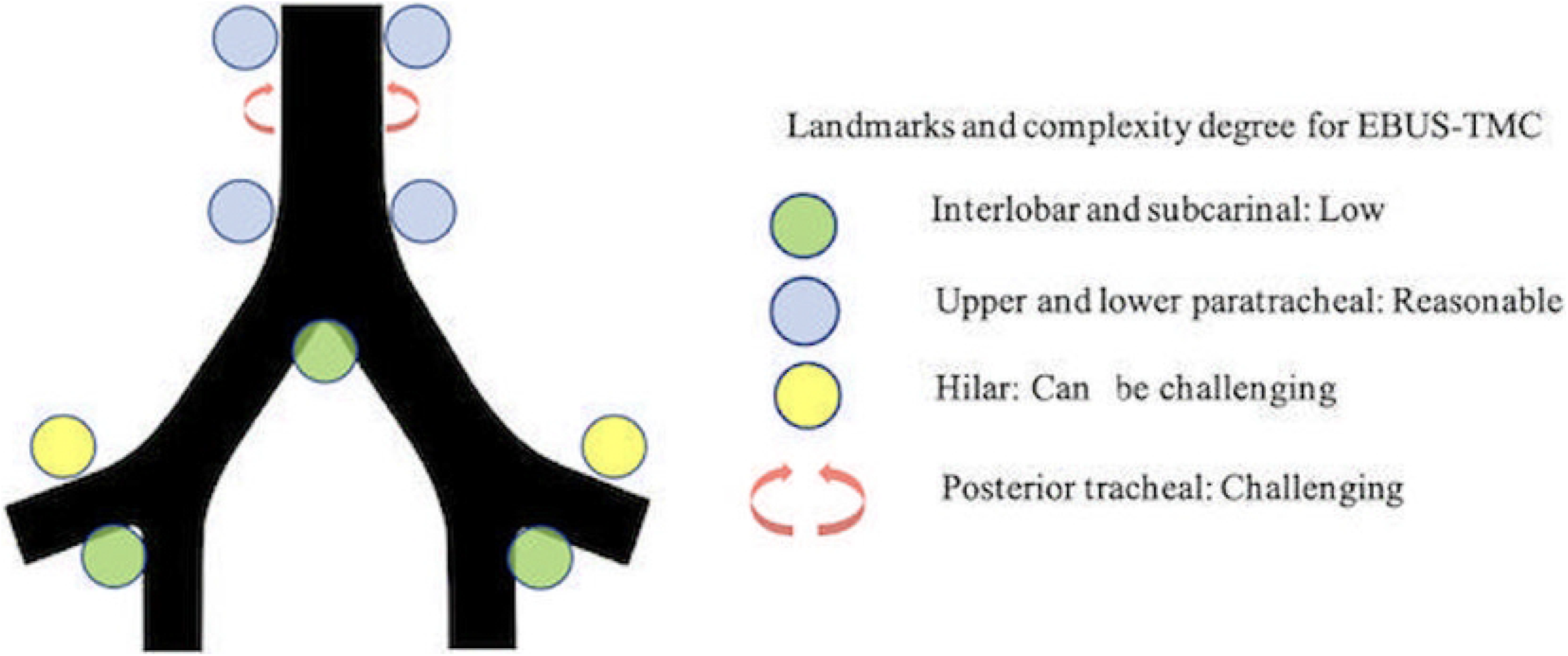
Landmarks and the complexity degree of performing EBUS-TBMC Abbreviation: Transbronchial mediastinal cryobiopsy guided by endobronchial ultrasound =EBUS-TBMC, Transbronchial needle aspiration =TBNA

### 4.2 Diagnostic yield

Various studies have demonstrated that incorporating EBUS-TBMC alongside conventional EBUS-TBNA can significantly improve the diagnostic yield, especially for benign disorders, lymphomas, and rare mediastinal conditions [5-6,8-9]. However, it is essential to acknowledge that intra-lesion necrosis or fibrotic lymph nodes may occasionally limit tissue availability, resulting in a potential reduction in diagnostic yield [10]. Another limitation of EBUS-TBMC involves addressing the heterogeneity of lymph nodes, as the procedure may not allow for biopsies at multiple sites, unlike EBUS-TBNA, which can sample a larger area through different needle entry points or the fanning technique [11].

Despite this limitation, EBUS-TBMC offers several advantages. Histology samples obtained from EBUS-TBMC enable more comprehensive testing compared to cytology examination, especially when dealing with poorly differentiated tumours. This aspect is particularly crucial for obtaining accurate diagnoses and guiding treatment decisions. One of the most significant clinical implications of EBUS-TBMC lies in its ability to provide sufficient tissue for complete molecular profiling before initiating targeted therapy for non-small cell lung carcinoma (NSCLC) [8]. Additionally, in cases of multiple synchronous/metachronous lung cancers, large tissue specimens are often required for comprehensive testing, underscoring the indispensability of EBUS-TBMC in such clinical scenarios.

In our study, we achieved an overall diagnostic yield of 83.3% (25 cases) with an 83.3% diagnostic yield for malignancy. All 12 patients diagnosed with NSCLC had adequate samples from EBUS-TBMC for next-generation sequencing testing. Nonetheless, it is essential to acknowledge that EBUS-TBMC did not establish a diagnosis in four cases. The contributing factors include poor patient tolerance, imprecise biopsy location, and extensive necrosis.

### 4.3 Complication

Minor bleeding was observed in six cases, but no intervention was required, indicating a manageable and self-limiting complication. To further mitigate the risk of bleeding, we recommend conducting Doppler assessments for vascular evaluation before the biopsy. This approach helps identify potential vascular structures and allows for safer probe placement during the procedure.

While rare, pneumothorax and pneumomediastinum have been reported in some cases [5]. As a precautionary measure, we routinely perform post-procedure chest radiographs to promptly detect any signs of air leak. Fortunately, no instances of pneumothorax, pneumomediastinum, mediastinitis, or mortality were observed in our cohort, which is consistent with previous studies demonstrating the overall safety of EBUS-TBMC [5-8].

In our study, we did not administer prophylactic antibiotics, even when sampling large necrotic lesions. This decision aligns with current guidelines and is supported by the lack of observed infections or complications related to the absence of antibiotic prophylaxis. Nevertheless, we emphasize the importance of cautious and skilled execution of EBUS-TBMC to minimize the risk of adverse events. While the procedure offers valuable benefits in obtaining larger tissue samples, operators should be well-trained and familiar with the technique’s intricacies to ensure patient safety.

### 4.4 Advantage and disadvantages

Despite some studies reporting mixed outcomes regarding procedure time [5-7], our observations suggest that EBUS-TBMC and EBUS-TBNA may have comparable overall procedure times. We have an ongoing study to validate this hypothesis. One significant drawback of EBUS-TBMC is its cost, necessitating careful consideration and justification of its use. The expense of a single-use cryoprobe is more than twice that of a TBNA needle, rendering an EBUS-TBMC procedure to cost almost three times more expensive. However, it is noteworthy that the future development of a cryo-needle may offer a potential solution to address this cost concern [12]. Table 2 provides an overview of the advantages and disadvantages of EBUS-TBMC. Notably, EBUS-TBMC has the potential to be advantageous in cases with low diagnostic yields using EBUS-TBNA, as it may help avoid repeat procedures, ultimately leading to cost savings. Further prospective studies are necessary to ascertain its efficacy.

**Table 2.**
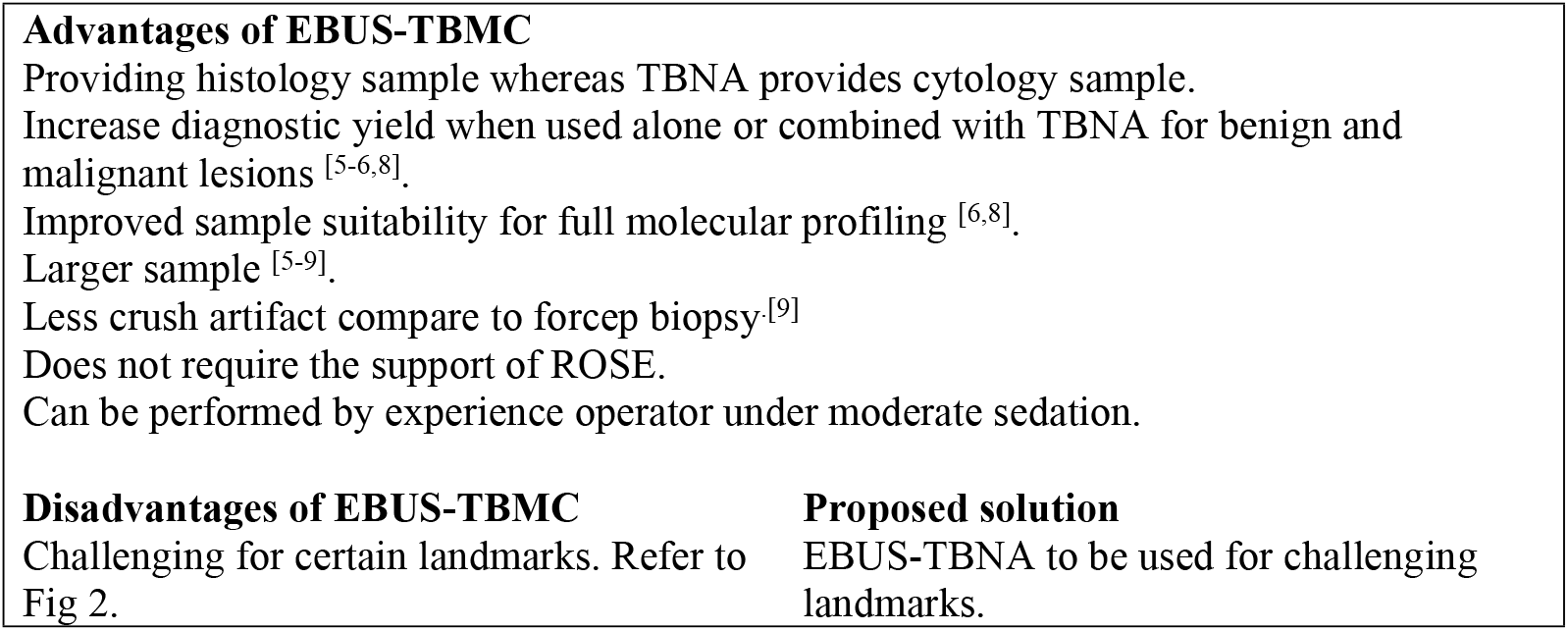
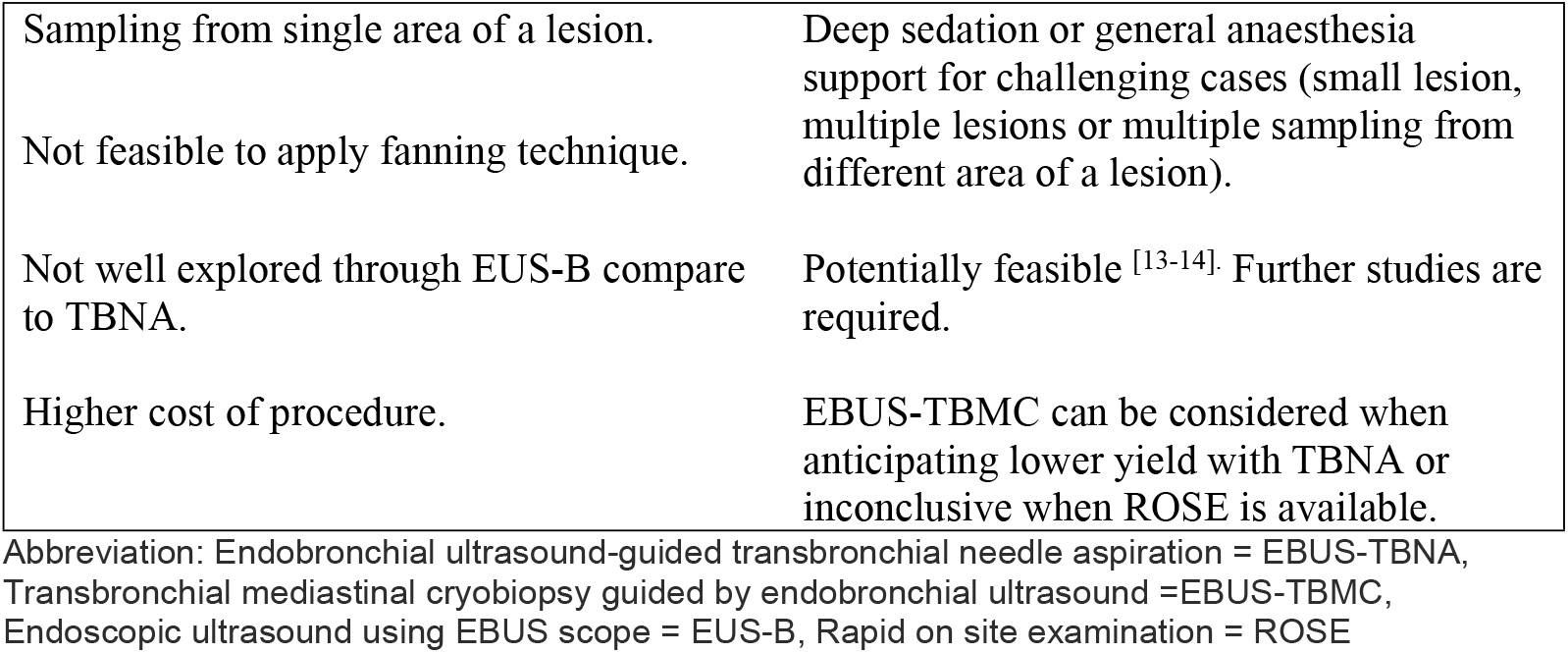
Advantages and disadvantages of EBUS-TBMC and the proposed solution.

### 4.5 Limitation of current study

The current study has several limitations that require consideration. One limitation of the study is the relatively small sample size, and the fact that it was conducted at a single center. As a result, the findings may not be fully generalizable to broader populations or diverse clinical settings. Conducting larger multi-center studies can help validate and enhance the robustness of the results. Another limitation arises from the retrospective nature of the study design, which may introduce potential biases such as selection bias, information bias, and recall bias. The reliance on existing data and historical records may impact the accuracy and completeness of the findings. Despite these limitations, the study provides valuable insights into the feasibility and utility of EBUS-TBMC. Recognizing and addressing these limitations in future research endeavours will contribute to a more comprehensive understanding of the clinical implications and benefits of EBUS-TBMC.

## 5. Conclusion

In conclusion, our study demonstrates the feasibility and safety of endobronchial ultrasound-guided transbronchial mediastinal cryobiopsy (EBUS-TBMC) through the biopsy track created by a TBNA needle. The procedure, performed under moderate sedation, provides a high diagnostic yield, offering valuable insights into mediastinal disorders and lung cancers. The favourable safety profile and larger tissue samples obtained through EBUS-TBMC enhance diagnostic accuracy and molecular testing, potentially optimizing therapeutic decision-making. As evidence continues to emerge, EBUS-TBMC has the potential to transition from an ancillary test to a primary method, complementing or even replacing EBUS-TBNA in certain clinical scenarios.

In summary, our study contributes valuable insights into the growing evidence on EBUS-TBMC. The procedure’s diagnostic capabilities, safety, and ease of implementation underscore its potential significance in diagnosing various mediastinal disorders and lung cancers. As the field advances, EBUS-TBMC holds promise as a key diagnostic modality shaping the future of diagnostic bronchoscopy.

## Data Availability

All relevant data are within the manuscript and its Supporting Information files.

